# User Preferences on Long-Acting Pre-Exposure Prophylaxis for HIV Prevention in Sub-Saharan Africa: A Scoping Review

**DOI:** 10.1101/2024.04.01.24305173

**Authors:** Brian Pfau, Arden Saravis BA, Sarah N. Cox, Linxuan Wu, Rachel Wittenauer, Emily Callen, Cory Arrouzet, Monisha Sharma

## Abstract

**Background:** Novel formulations for PrEP such as injectables, implants, and intravaginal rings are emerging as long-acting alternatives to daily pills for the prevention of HIV. Sub-Saharan Africa has the highest HIV burden as well as the highest PrEP coverage globally. As long-acting formulations continue to become available, it is crucial to understand the product preferences of potential users.

**Objective:** To conduct a scoping review focused on the region of Sub-Saharan Africa to understand which PrEP products, especially long-acting formulations, different patients and demographic groups prefer as well as the factors that influence their preferences.

**Design:** We identified 34 publications published between 2014 and 2024 that assessed preferences regarding at least one long-acting PrEP product in the region of Sub-Saharan Africa.

**Results:** Participants preferred longer-acting products over oral pills when given the choice in almost all studies. On-demand PrEP was commonly preferred over daily dosing, and long-acting products were preferred over on-demand dosing. A majority of studies found injectables to be most commonly preferred compared to daily oral PrEP, implants, and rings. This preference was observed across a range of demographics including men and women, youth, men who have sex with men, and female sex workers. Duration, efficacy, and discretion were the three most important factors influencing participants’ choices.

**Conclusions:** Long-acting products, especially injectables, are acceptable for a wide range of individuals at risk of HIV in Sub-Saharan Africa and tend to be preferred over daily oral pills. Participants expressed a diversity of values and opinions regarding preferences, emphasizing the benefit of providing multiple formulations to maximize coverage over heterogeneous populations.

**Strength and Limitations of this Study:** Some key populations, such as transgender women, were underrepresented in the literature. With most studies published before long-acting products became widely available, the hypothetical preferences of non-experienced users may differ from preferences in practice.

## INTRODUCTION

Pre-exposure prophylaxis (PrEP) is a promising tool in the HIV prevention portfolio and is highly effective when used with high adherence. Although PrEP uptake has increased significantly since its introduction in 2012, coverage remains low among people at HIV risk, with 1.6 million estimated global users–well below the United Nations target of 10 million users by 2025 [1]. Additionally, PrEP retention and adherence among those who initiate is suboptimal, reducing its effectiveness [2]. Barriers to PrEP uptake and adherence include pill burden, stigma, and lack of discretion associated with oral tablets. In light of these challenges, research has focused on developing several long-acting PrEP formulations, including injectables, implants, and vaginal rings, which could increase PrEP coverage and adherence by providing more convenient and discreet options. Cabotegravir, a bimonthly antiretroviral, was the first long-acting injectable (LAI) approved for clinical use by the US Food and Drug Administration in December 2021. The intravaginal dapivirine ring (DVR), developed primarily for women in low-income countries, was recommended for use by the World Health Organization (WHO) in January 2021 [3]. An implant with six-month duration is currently in development. Finally, lenacapavir is a first-in-class twice-yearly injectable which was approved for treatment of HIV in 2022 and is currently undergoing Phase III trials for use as PrEP as well. In addition to long-acting formulations, oral on-demand PrEP (or event dosing) is an alternative dosing schedule with comparable efficacy to daily use in which patients take a double dose up to two hours before a potential exposure and then once every 24 hours the following two days.

Understanding preferences for PrEP products among subgroups at risk of HIV infection is crucial for maximizing PrEP coverage and impact. The aim of this scoping review was to synthesize the literature on PrEP perceptions and preferences in Sub-Saharan Africa (SSA), the region most impacted by the HIV epidemic. PrEP is a priority intervention for scale-up in SSA and policymakers must decide which products to implement and how to tailor demand generation strategies. Understanding the relative preference of LA PrEP compared to oral PrEP by subpopulation is also important for commodity planning.

## METHODS

We conducted a scoping review of peer-reviewed literature on preferences for long-acting PrEP from 2014 to 2024. We adopted a systematic approach, following Preferred Reporting Items for Systematic Reviews and Meta-Analyses extension for Scoping Reviews (PRISMA-ScR) guidelines [4]. We conducted a keyword search for relevant articles first on PubMed followed by Google Scholar, which returned the same relevant publications as PubMed. We included the following keywords during our search: “PrEP”, “long-acting”, “discrete choice”, “preferences”, “Africa”, “on demand” (Table S1). We also received grey literature and articles not yet published from a research collaborator.

### Inclusion and exclusion criteria

Studies were eligible for inclusion if they met the following criteria: (a) original research; (b) peer-reviewed and published in English between 2014 and 2024 or provided by a research partner; (c) research conducted in SSA; (d) evaluating preferences for at least one long-acting or on-demand PrEP product alongside daily oral PrEP.

### Data screening

Two reviewers (BP and AS) screened the list of references for inclusion into the study. First, titles and abstracts of articles were reviewed and then selected articles underwent full text review. Disagreements were resolved via team discussions. We extracted data on title, publication year, location of data collection, population assessed, main findings, study strengths and limitations. We categorized each study by region and demographic focus.

### Quality assessment

One reviewer (BP) conducted a quality assessment of the 34 eligible studies based on generalizability to the target population, participant acceptance rate, and PrEP experience/naïveté of the sample (Table S2).

## RESULTS

### Characteristics of studies

Of 214 unique citations identified, 35 articles met eligibility criteria and were included in the review (Figure 1). Study characteristics are summarised in Table 1. Most studies were cross-sectional (22) including nine discrete choice experiments (DCEs). Most or all individuals in 11 studies had prior experience with oral PrEP, while nine studies were conducted among exclusively PrEP-naïve participants (Table S2). 10 studies were conducted among individuals participating in randomized clinical trials for PrEP [5–11], cohort studies [12,13], or PrEP implementation projects [14].

**Figure 1.**
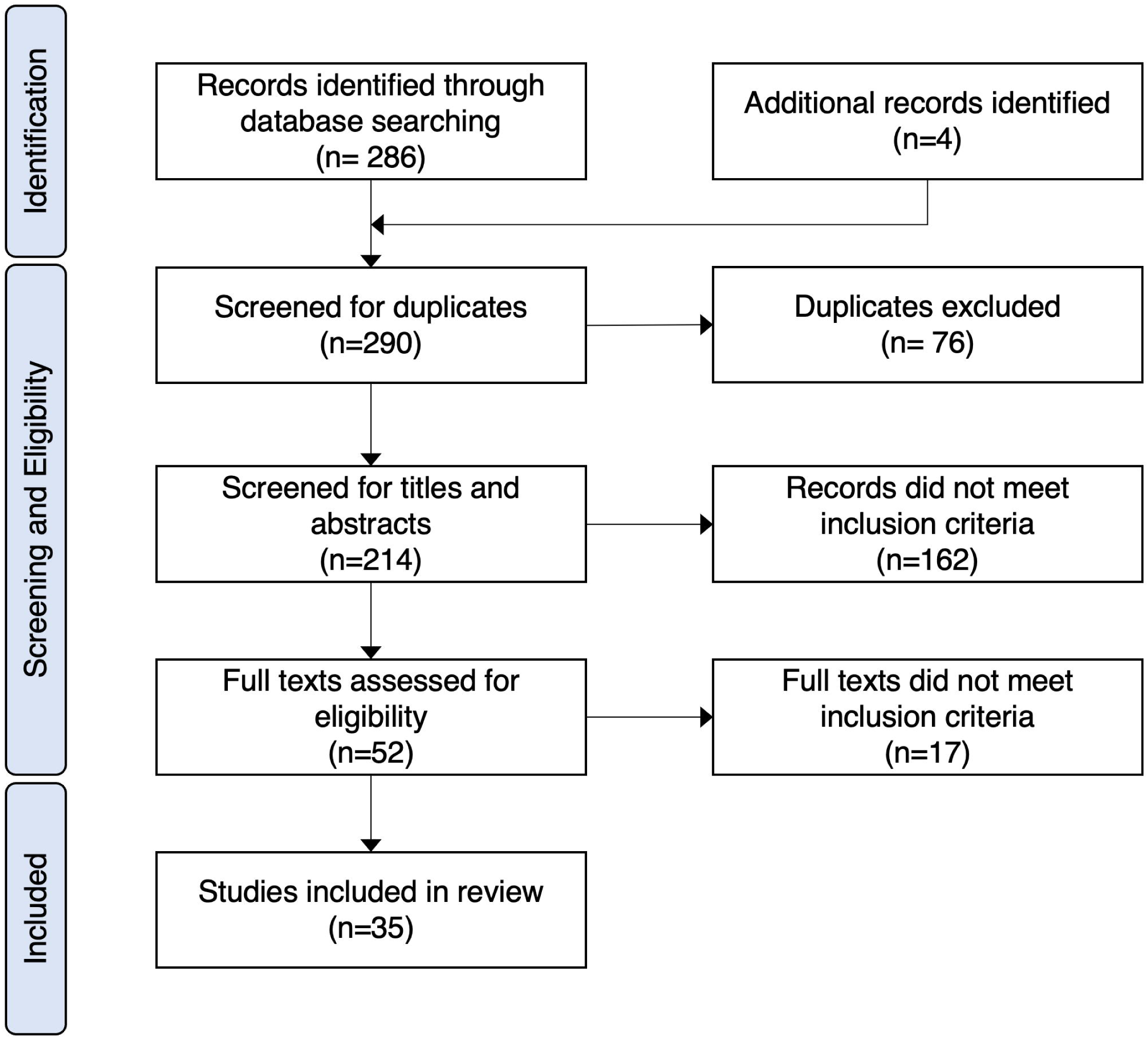
PRISMA flow diagram

**Table 1.** Characteristics of studies.

| Characteristics | N of studies (%) |
| --- | --- |
| **Formulation** |  |
| Long-acting Injectable (LAI) | 26 (74%) |
| Intravaginal dapivirine ring (DVR) | 14 (40%) |
| Implant | 18 (51%) |
| On-demand | 4 (11%) |
| Other | 10 (29%) |
| **Demographic** |  |
| Men | 19 (54%) |
| ABYM | 12 (34%) |
| MSM<sup>1</sup> | 3 (9%) |
| Heterosexual men<sup>1</sup> | 2 (6%) |
| Women | 29 (83%) |
| AGYW | 17 (49%) |
| FSW | 5 (14%) |
| Transgender women<sup>1</sup> | 1 (3%) |
| Women only | 15 (43%) |
| Providers | 1 (3%) |
| **Location** |  |
| South Africa | 25 (71%) |
| Kenya | 9 (26%) |
| Uganda | 8 (23%) |
| Zimbabwe | 8 (23%) |
| Tanzania | 2 (6%) |
| Eswatini | 2 (6%) |
| Malawi | 2 (6%) |
| Nigeria | 1 (3%) |
| US | 1 (3%) |
| **Study Design** |  |
| Randomized controlled trial | 6 (17%) |
| Discrete choice | 9 (26%) |
| Cross-sectional | 13 (37%) |
| In-depth Interview | 10 (29%) |
| Focus group discussion | 10 (29%) |
| **Publication Type** |  |
| Peer-reviewed | 32 (91%) |
| Grey literature | 3 (9%) |

<sup>1</sup>Study sought to examine this group explicitly.

Studies included both men (n=19) and women (n=29), including 15 studies which focused on women’s preferences specifically. Five studies focused on males specifically, including three studies assessing preferences of men who have sex with men (MSM) [15–17], and two among heterosexual men [5,18]. Transgender women were included in only one study alongside MSM [17]. A majority of studies (n=19) included or focused on youth, here referred to as adolescent boys and young men (ABYM) or adolescent girls and young women (AGYW). Female sex workers (FSWs) were assessed in five studies.

South Africa was the most represented country (n=25), followed by Kenya (n=9), Zimbabwe (n=8) and Uganda (n=8). Seven papers presented findings from three countries combined (Uganda, Zimbabwe, and South Africa). Other papers included Tanzania (n=2), Eswatini (n=2), Malawi (n=1), and Nigeria (n=1).

The majority of studies evaluated LAIs (26), followed by implants (n=18), the DVR (n=14), and other prevention methods such as condoms, films, or gels (n=10). All formulations were compared against daily oral PrEP. Three studies compared preferences between daily oral PrEP and oral on-demand dosing, and one study compared on-demand dosing to injectables and implants. Table 2 summarises the results of the studies included.

**Table 2.** Summary of studies.

| Author et al. (Year) | Demographic | Location | Study Design | Formulation | Sample size | Main findings | Ref |
| --- | --- | --- | --- | --- | --- | --- | --- |
| Mack et. al. (2014) | Women, AGYW, FSW | South Africa, Kenya | Focus group | LAI, Other | 101 | 1. The majority of FSWs in Kenya preferred LAIs over oral pills or vaginal gels for the convenience of long-term protection and the perception of injections as most discreet. By contrast, few were interested in a gel. Many were interested in the prospect of accepting condomless sex for more money while being protected by PrEP.<br>2. Preferences among AGYW were equivocal, with some perceiving pills as safer than injections while others perceived LAIs as safer as well as more convenient, long-lasting, and discreet.<br>3. Members of serodiscordant partnerships strongly preferred LAIs for duration and convenience, though some were hesitant due to fear of needles or injection pain. | [27] |
| Luecke et al. (2016) | Women | South Africa, Uganda, Zimbabwe | Interview | DVR, LAI, Implant, Other | 68 | 1. 81% of women preferred long-acting products, citing duration, safety, ease of use, partner concerns, and route of administration.<br>2. One quarter of women expressed concern about being perceived as HIV-positive if others knew they were taking pills every day. | [6] |
| Van der Straten et al. (2017) | Women | South Africa, Uganda, Zimbabwe | RCT, Interview | DVR, LAI, Implant, Other | 71 | 1. When women could select more than one product as “most preferred,” the ring was selected by 94% of women compared with 39% for implants and 33% for LAIs.<br>2. Participants broadly appreciated continuous protection, discretion, and peace of mind arising from simplified use and infrequent dosing versus worry about forgetting daily doses.<br>3. Women expressed concern with several formulations about negative reactions from a male partner if he discovered they were using the PrEP product, e.g. feeling the ring, noticing the implant under the skin, or lubrication from the other vaginal formulations. | [11] |
| Krogstad et al. (2018) | Men, ABYM, Women, AGYW | South Africa | Focus group | Implant | 105 | 1. In a focus group for the design of a long-acting implant, longer duration ($\geq 6$ months) was a significant attribute among interviewees.<br>2. Discreetness and comfort were valued with desire for a flexible vs stiff implant.<br>3. Biodegradability desired by almost all participants to avoid removal and associated clinic visits. | [30] |
| Siedner et al. (2018) | ABYM, Women | Eswatini | Discrete choice | DVR, LAI, Implant | 109 | 1. 75% preferred a two-month LAI over oral daily. This preference held across sexes, age, education, and sexual behavior. | [48] |
| Van der Straten et al. (2018) | Women, AGYW | South Africa, Kenya | RCT, Cross-sectional | DVR, LAI, Other | 249 | 1. Formulations most preferred by AGYW were LAIs (62%), followed by oral pills (15%), DVR (12%) and condoms (10%). The most frequently least preferred formulations were the DVR (41%) and oral pills (35%).<br>2. Between the two countries, South African women had twice the odds of choosing LAIs, while Kenyan women had twice the odds of choosing oral pills. | [47] |
| Cheng et al. (2019) | Heterosexual men | South Africa | Discrete choice | LAI, Other | 178 | 1. Among heterosexual men, 48% preferred LAI versus oral (33%) and condoms (20%).<br>2. Men with children and men who were less risk averse were more likely to prefer LAI.<br>3. Participants concerned with high risk of STIs other than HIV tended to prefer other methods. | [18] |
| Harling et al. (2019) | Women | Tanzania | Cross-sectional | DVR, LAI, Other | 66 | 1. While only 5% of respondents had initially heard of PrEP, 79% were somewhat/very interested in LAI, compared to 54% interested in oral daily, 38% in vaginal gel, and 11% in DVR.<br>2. LAI was ranked most preferred among the four modalities, while 82% picked oral daily as either first or second preferred. | [19] |
| Krogstad et al. (2019) | Providers | South Africa | Interview | LAI, Implant | 30 | 1. Healthcare providers asked for input on a design for a PrEP implant preferred a product that is long-lasting (>6 months), biodegradable to avoid need for removal, and a flexible while still palpable design.<br>2. Providers cited understaffed clinics and inadequate training for contraceptive removal as major factors influencing their preferences. | [31] |
| Montgomery et al. (2019) | Men, ABYM, Women, AGYW | South Africa | Interview, Focus group | DVR, LAI, Implant | 95 | 1. Majority of participants interviewed preferred LAI or implants. Many expressed their major influencing factor as efficacy, even if the formulation was expected to be painful.<br>2. Oral PrEP-experienced men often reported difficulty with the daily dosing regimen, especially on weekends. Some oral PrEP-experienced women felt the pills were more discreet since they do not leave a mark like an injection.<br>3. Vaginal ring PrEP-experienced women expressed a similar focus on efficacy and expressed concern about discomfort during sex and whether their partner would be able to feel the ring.<br>4. LAI PrEP-experienced women appreciated the longer dosing frequency and cited side effects as chief complaint, though they perceived the injection to have greater efficacy than other methods. | [7] |
| Tolley et al. (2019) | Women | South Africa, Zimbabwe, US | RCT, Cross-sectional, Focus group | LAI | 136 (100 African) | 1. 93% of non-U.S. participants preferred LAI compared to 64% of U.S. participants.<br>2. Participants liked the idea that PrEP could be easier to use and of longer duration, though about a third of participants expressed concerns about potential side effects and pain. | [10] |
| Kidman et al. (2020) | Men, ABYM, Women, AGYW | Malawi | Cross-sectional | LAI | 2089 | 1. Among 10-16-year-old youth, 80% expressed willingness both to use oral and injectables, but only 52% of girls and 48% of boys would still consider using PrEP if there were side effects.<br>2. 87% of caregivers indicated that they would want their child to take a daily pill to prevent HIV. | [38] |
| Laher et al. (2020) | Men, ABYM, Women, AGYW | South Africa | Focus group | DVR, LAI, Implant, Other | 68 | 1. In focus groups, participants expressed preference for long-lasting duration, favoring LAIs and implants for their efficacy, discretion, and duration.<br>2. Men and women mentioned the difficulty of swallowing large pills as well as concern that others might assume they had HIV if they saw them taking daily pills. | [29] |
| Minnis et al. (2020) | Men, ABYM, Women, AGYW | South Africa | Discrete choice | LAI, Implant | 807 | 1. Strong preference for less frequent dosing and for injectables over implants among male and female urban youth including MSM.<br>2. Participants were willing to trade their preferred formulation for one with a longer dosing frequency. | [23] |
| Van der Straten et al. (2020) | Men, Women | South Africa, Uganda, Zimbabwe, Malawi | Focus group | DVR | 128 | 1. Pregnant and breastfeeding women were overall accepting of PrEP and highly valued safety for mother and child regarding PrEP use. They stressed the importance of personal choice in HIV prevention and wished to have a variety of options so they could choose the product that worked best for them.<br>2. Participants feared exacerbation of pregnancy symptoms such as vaginal discomfort with the DVR or nausea for oral pills. Taboos against vaginal insertion or taking medicine during pregnancy could be a barrier to initiation for both formulations.<br>3. Partner support was paramount, and women typically considered PrEP use to be a decision made in conjunction with their partners. | [25] |
| Montgomery et al. (2021) | Men, ABYM | South Africa | Discrete choice | LAI | 406 | 1. Young men reported several features to be "very important": perceived efficacy (94%), where one has to go to get it (88%), dosing frequency (87%), and removability in the event of side effects (85%).<br>2. Both MSW and MSM valued privacy and discretion, though more MSW valued being able to use PrEP without their partner knowing (46%) | [41] |
|  |  |  |  |  |  | compared to MSM (27%).<br>3. 94% reported willingness to pay for an LAI. |  |
| Ngure et al. (2021) | Women, AGYW | Kenya | RCT, Cross-sectional | LAI | 350 | 1. Among young women exiting the MPYA PrEP monitoring study, 36% preferred injectable Cab-LA, 34% preferred daily pills, 22% preferred implants, 15% preferred vaginal rings.<br>2. No association between preference and age, HIV risk, marital status, contraceptive method among those using contraception, or other variables. | [8] |
| Beckham et al. (2022) | Women, FSW | Tanzania | Cross-sectional, Interview, Focus group | DVR, LAI | 496; 10; 20 | 1. Where 92% of FSWs participating were initially unaware of PrEP, 88% preferred LAI over oral daily, citing dosing frequency, discretion, and belief in higher efficacy.<br>2. 58% felt PrEP was personally worth it to take, and those who did were more likely to have recent STI symptoms or diagnosis.<br>3. Many FSWs stressed that they would not reduce condom use if they took PrEP. | [26] |
| Brown et al. (2022) | Women, AGYW, FSW | South Africa | Interview | Implant | 36 | 1. In qualitative interviews, end-users generally perceived some drawbacks of the implant such as side effects and pain during insertion but believed these were outweighed by the benefits of a highly effective and long-lasting PrEP product.<br>2. HCPs acknowledged concerns about increased risky sexual behavior but recognized value in the implant's extended protection and improved adherence compared to oral PrEP.<br>3. Despite low awareness of oral PrEP, end-users expressed willingness to try a PrEP implant. FSWs liked the ability to protect themselves from HIV without requiring a client to use a condom. | [20] |
| Dietrich et al. (2022) | Men, ABYM, Women, AGYW | South Africa, Uganda, Zimbabwe | Cross-sectional | On-demand | 1330 | 1. 60% stated a preference for on-demand, with males and older young adults more likely to prefer on-demand.<br>2. Preference for on-demand PrEP decreased with more frequent sexual activity in the last month. | [32] |
| Little et al. (2022) | Women, AGYW, FSW | South Africa | Discrete choice | Implant | 600 | 1. 78% of respondents stated they would be likely or very likely to use an implant if one were available. 82% expressed preference for a dual use product for PrEP and contraception.<br>2. Broad preference for 24-month vs 6-month protection interval and for a biodegradable (dissolvable) product. | [21] |
| Mayanja et al. (2022) | Women, AGYW | Uganda | Discrete choice | DVR, LAI, Implant, Other | 285 | 1. 47.6% preferred oral PrEP, 52.4% preferred hypothetical PrEP alternatives.<br>2. Preference for oral PrEP was associated with 50% higher PrEP uptake.<br>3. Low awareness of oral PrEP (24.5%), and even lower awareness of hypothetical alternatives (LAI 4.2%, DVR 2.3%, HIV vaccine 1.5%, implant 0%). | [13] |
| Ogunbajo et al. (2022) | Men, MSM | Nigeria | Cross-sectional | LAI, Implant, Other | 305 | 1. 88% of MSM, were willing to use LAI with 44% of participants preferring it. 21% preferred daily oral, 17% preferred lubricants, 10% preferred all formulations equally, while only 6% preferred implants.<br>2. Those who preferred LAI were more likely to be single, report inconsistent condom use, and report having a primary care provider. | [15] |
| Webb et al. (2022) | Men, ABYM, Women, AGYW | South Africa, Uganda, Zimbabwe | Cross-sectional | On-demand | 1330 | 1. PTSD symptoms were not associated with willingness to take PrEP or preference for on-demand vs. daily PrEP. | [33] |
| Bailey et al. (2023) | Men, Women, MSM, Transgender women | Kenya | Interview | LAI, Implant, Other | 423 | 1. In pairwise comparisons, quarterly injections were most preferred (26%), followed by monthly pills (23%), a yearly implant (19%), condoms (12%), and oral daily (1%).<br>2. When “forced” to choose the most preferred product, 37.1% preferred a quarterly injection, 34.8% preferred a monthly pill, 25.8% preferred a yearly implant, and 2.4% preferred oral daily.<br>3. Transgender women were more likely to prefer the implant over quarterly injections than gay or bisexual men.<br>4. Muslim participants had considerably greater preference for implants compared to Christians, adherents of African indigenous religions, and religiously unaffiliated participants. | [17] |
| Jansen van Vuuren et al. (2023) | Women | South Africa | Interview | LAI, Implant | 425 | 1. Women were most likely to prefer the same PrEP formulation as that which they were using or had used for contraception. | [14] |
| Kakande et al. (2023) | Men, ABYM, Heterosexual men | South Africa, Uganda, Zimbabwe | RCT, Cross-sectional, Interview | On-demand | 647 | 1. 65.2% of participants stated a preference for on-demand PrEP. Preference was higher in Uganda (76.8%) and Zimbabwe (70.4%) than in South Africa (45.5%).<br>2. Preference for on-demand PrEP increased with age.<br>3. Reasons for preferring on-demand PrEP included not liking daily tablets (38%), not thinking one is exposed to HIV frequently enough to warrant a daily pill (17%), fear that taking daily PrEP might make others think one has HIV (15%), and concern for pill fatigue (11%).<br>4. Reasons for preferring daily PrEP included continuous protection (45%), better protection overall (19%), and ease of daily routine vs. remembering to take pill before having sex (15%). | [5] |
| Little et al. (2023) | Women, AGYW | South Africa, Kenya, Eswatini | Discrete choice | LAI, Implant | 1263 | 1. Women most preferred 12-18-month removable implants followed closely by 3-6-month LAIs, with daily oral pills, weekly skin patches, and the monthly DVR ranked lowest preferred. There was a strong preference for multi-purpose products which could prevent pregnancy and/or STIs.<br>2. Efficacy, duration, and reversibility were the most important attributes for implants. | [22] |
| Mataboge et al. (2023) | Men, ABYM, Women, AGYW, MSM, FSW | South Africa | Focus group | DVR, LAI | 109 | 1. Participants overall preferred LAI (Cabotegravir) over DVR. Most cited reduced time spent at overcrowded clinics, ease of method continuation, and removed burden of daily dosing as reasons for embracing long-acting formulations.<br>2. Most AGYW and all pregnant AGYW and FSWs would not use the DVR because of perceived side effects, and some ABYM worried about their partner's potential pain during intercourse and lower efficacy. Participants cited familiarity with injectable contraceptives as part of the acceptability of LAIs. However, some AGYW feared similar side-effects to injectable contraceptives, e.g. menstruation cessation and weight gain. | [16] |
| Ngure et al. (2023) | Women, AGYW | South Africa, Uganda, Zimbabwe | RCT, Cross-sectional | DVR | 247 | 1. Similar proportions of preference between the ring (38.1%) and oral PrEP (40.5%), with 19% preferring both equally. | [9] |
| Tran et al. (2023) | Men, Women | Kenya | Discrete choice | LAI | 50 | 1. HIV+ participants generally preferred hypothetical long-acting formulations to their current ART therapies. | [49] |
| Wara et al. (2023) | Women | South Africa, Kenya | Discrete choice | DVR, LAI | 394 | 1. 75% of participants preferred a potential LAI over daily oral PrEP.<br>2. In South Africa, longer duration of effectiveness was the major factor influencing preference (87% South Africa vs. 42% Kenya). Discretion was a larger factor in Kenya (5% South Africa vs. 49% Kenya).<br>3. Conversely, 87% of participants preferred oral PrEP over a vaginal ring mostly due to concern about discomfort (82% South Africa, 48% Kenya).<br>4. Preferred frequency of PrEP use by ranking order was once a year (31%), once a month (16%), once every 2-3 months (15%), before sex (13%), every day (12%) and once every six months (11%). | [12] |
| Mthimkhulu et al. (2024) | Men, ABYM | South Africa | Cross-sectional, Focus group | On-demand, Implant | 145 | 1. Most men would consider using a monthly pill (74.6%), the implant (62.7%), oral on-demand (59.2%), and the six-month LAI (57.7%). 50% would consider oral daily, and 43.7% would consider a bimonthly LAI. However, in group interviews men generally agreed that any product would be acceptable as long as it was effective.<br>2. If only one choice were available, the most preferred options were a monthly pill (31.7%), the six-month LAI (28.2%), and the implant (19.7%).<br>3. Side effects were the primary concern around PrEP, especially sexual/fertility issues and discomfort on implant insertion. | [34] |
| Gates Foundation [not published] | Women | South Africa, Kenya | Focus group | Implant | 32 | 1. In an assessment of potential combination PrEP and contraceptive methods, implants were perceived as invasive and too visible to be discreet, and other options were perceived as more convenient.<br>2. Separate coadministered LAI and contraceptive injections were acceptable as long as both injections were of the same duration. | [50] |
| Were [not published] | Men, Women | Kenya | Cross-sectional, Focus group | DVR, LAI, Implant | 6013; 257 | 1. 91.4% of participants willing to use LAI preferred a six-month duration over a two-month. 63.6% preferred subcutaneous injection over intramuscular injection and 92.4% preferred provider-administration over self-administration.<br>2. 41.5% of participants willing to use implants preferred a biodegradable that did not need to be removed, while 58.5% preferred a nonbiodegradable because it could be removed if needed and/or because they feared its absorption into the body.<br>3. Among those who declined PrEP, most cited reasons were the perceived burden of taking a daily pill (35.4%), fear of side effects (21.2%), already consistently using condoms (9.9%) and trusting their partner (9.5%).<br>4. Participants most desired to use PrEP if alternative formulations were available without side effects or the burden of a daily pill and its associated stigma. | [24] |

### Preferred products by subpopulation

#### Women

Adult women and adolescent girls and young women (AGYW) most commonly preferred LAIs over other long-acting products as they were perceived as having greater efficacy and favorable duration, and as suitably discreet. Women participating in the CAPRISA 082 study in South Africa who had previous experience using an implantable or injectable contraceptive method were more likely to choose that method for PrEP [14]. In a South African focus group, despite overall preference for LAIs over oral pills and the DVR, a few AGYW were dissuaded from injectable PrEP for fear of side effects similar to injectable contraceptives, such as menstruation cessation and weight gain [16].

Interest in the DVR was mixed. The ring was regarded positively among participants in the ASPIRE study, a multinational clinical trial of the DVR. Where women could select more than one product as “most preferred,” 94% of women selected the ring, compared with 39% for implants and 33% for LAIs [11]. However, many women expressed concern about discretion, such as whether their partner could feel the ring during intercourse or whether it could come out accidentally [7]. In most studies, DVR was less frequently preferred compared to LAIs [7,12,16,19]. Most AGYW in one focus group would not use the DVR due to perceived side effects, especially pain during intercourse, as well as concern for hygiene and use during menstruation [16].

Women reported broad interest in using implantable PrEP and stated that the benefits of an effective and long-lasting product outweighed potential drawbacks such as side effects or pain on insertion. They also expressed interest in a dual use product for PrEP and contraception in surveys and interviews. Since implants are already a common modality for extended-release contraceptives, a device which combines the two could be attractive to women seeking protection from both HIV and pregnancy. [7,15,17,20–24].

In a study of pregnant and postpartum women participating in the PrEP-PP and PrIMA-X trials in South Africa and Kenya, respectively, many voiced safety concerns for the mother and infant during pregnancy and breastfeeding. These women, most of whom had recent experience with oral PrEP, tended to prefer long-acting products over daily oral for their longer duration (especially in South Africa) and increased discretion (especially in Kenya) [12]. All pregnant AGYW interviewed in a study in South Africa would not use the DVR on account of perceived side effects [16]. Pregnant and postpartum women in multi-national focus groups mentioned fears of side effects ranging from exacerbation of pregnancy discomfort to severe outcomes such as miscarriage or birth defects. However, they were overall accepting of PrEP as long as it was safe and effective, and stressed the importance of choice and the ability to choose the formulation that worked best for them [25].

#### Female sex workers (FSWs)

Awareness of PrEP was low among FSWs in South Africa [20] and Tanzania [26], but after learning about PrEP most participants were willing to use it, particularly those who had recent symptoms or diagnosis of a sexually transmitted infection (STI). Many FSWs stated that protection by PrEP could enable them to have sex with clients without requiring condoms [20] or to accept condomless sex at a higher price [27], highlighting a potential concern regarding risk compensation. However, others viewed PrEP as a protective complement to condoms and stated that they would continue to use them to prevent STIs and pregnancy [26]. 86% of FSWs responded that they would likely use an implant in a South African DCE [21]. This high proportion of acceptability was complemented by qualitative interviews in South Africa [20], where FSWs commended the benefit of continuous protection and noted that it would be worth a brief amount of pain during insertion. Similarly, 88% of Tanzanian sex workers preferred LAIs over oral daily, although a smaller majority (58%) felt PrEP was generally worth taking. LAIs were considerably preferred over the DVR in interviews with South African FSWs; all sex workers stated they would not use the DVR on account of perceived side effects and concern that a client might notice the device [16].

#### Men

MSM tended to prefer LAIs over implants or oral pills. Among Nigerian MSM, preference for LAIs was associated with single relationship status, inconsistent condom use, and having a primary care provider [15]. Duration/dosing frequency was a highly prioritized product attribute for male participants. Oral PrEP-experienced young men in South Africa reported difficulty with daily dosing [7]. Men of all orientations valued privacy and discretion, though more men who have sex with women (MSW) valued being able to use PrEP without their partner knowing compared to MSM [28].

Preferences among heterosexual men were specifically assessed in two studies [5,18]. In a survey of urban heterosexual men in South Africa, 48% preferred LAIs compared to 33% who preferred oral and 20% who preferred condoms alone. Men who had children or who were less risk-averse were more likely to prefer LAIs. Men concerned with high risk of STIs other than HIV were more likely to prefer condoms over LAIs alone. As choices were discretely ranked, it was not clear how many men who preferred condoms for their protection against other STIs would prefer to use LAIs and condoms in combination. [18] In a mixed methods study conducted in South Africa, Uganda, and Zimbabwe among participants in the CHAPS trial, a majority of heterosexual male youth (65%) preferred on-demand oral PrEP compared to daily dosing. Those who did not believe they were exposed to HIV regularly enough to warrant taking a daily pill tended to prefer on-demand dosing in qualitative interviews. [5]

### Product attributes driving preference

#### Duration

Product duration was the most important factor driving user preferences in many studies across demographics [7,10,12,23,28]. Oral PrEP-experienced pregnant and postpartum women most commonly cited product duration as a factor for switching to LAI [12]. Oral PrEP-experienced men also frequently reported difficulty adhering to a daily dosing schedule, especially on weekends, as well as difficulty swallowing the pill itself [7]. (Note that the most common oral PrEP formulation, emtricitabine-tenofovir, is a very large tablet [19 mm] which can be difficult to swallow even for users who take other, smaller tablet medications.) Male and female youth in a South African DCE valued product duration highly and were typically willing to trade their preferred product for one with a longer dosing frequency [23].

Notably, some youth in interviews in South Africa suggested an ideal formulation as a monthly rather than a daily pill [16]. This would reduce the burden of daily dosing for users who preferred an oral tablet over other long-acting formulations, such as pain associated with injections.

#### Efficacy

Many participants emphasized PrEP efficacy as an important determinant of product preference. They noted that efficacy was tied to longer duration due to the difficulty of adhering to a consistent dosing schedule, which hinders the observed effectiveness of oral PrEP. PrEP effectiveness was the strongest factor driving preference in several studies, notably among pregnant and postpartum women [12] and among youth populations [7,28]. High PrEP effectiveness outweighed participant concerns about side effects such as pain upon injection or product insertion.

#### Discretion

Discretion (i.e., being able to use a PrEP formulation without a partner or the community knowing) was a commonly mentioned concern, particularly among women and persons who did not wish to disclose their PrEP use to their partners. In several studies evaluating oral PrEP alongside LA formulations, including women [11], FSWs [26], and adults [24], participants expressed concern about stigma associated with daily pill use. They feared that others would think they had HIV if they discovered that they were taking a daily antiretroviral pill. Similarly, a visible preventive product could be seen as a mark of sexual indiscretion or promiscuity, as voiced in one South African focus group including participants of all genders and sexual orientations [29]. According to them, using PrEP could sow distrust or signal infidelity among users with romantic partners, and a woman who used PrEP might be seen as sexually promiscuous.

Notably, product discretion was described differently by different individuals. While many participants found daily pills indiscreet, others felt oral PrEP was the most discreet since it does not leave a mark like an injection [7]. Others, especially women, were concerned about their partner knowing about their PrEP use. Many participants expressed disinterest in the DVR for fear that their partner would feel the ring during intercourse, or that it might fall out and cause embarrassment. Concerns about a partner noticing signs of PrEP use were also mentioned relating to the implant; many participants in interviews disliked the notion of a visible device under the skin and preferred one that would not be seen by others.

#### Implant biodegradability

In general, participants preferred biodegradable implants which would not need to be removed by a provider, alleviating the need for an extra clinic visit and pain during removal [21,30]. South African healthcare providers also stressed this feature in qualitative interviews [31]. However, in one study in Kenya, many participants (especially FSWs) preferred a nonbiodegradable implant for its reversibility, as it could be removed if needed. Some also expressed fear about the effects of the degraded materials being absorbed into the body [24].

#### Logistical challenges

Long-acting formulations addressed the challenges of frequent visits to clinics or pharmacies, which participants variously described as overcrowded, lacking in privacy, and inconvenient or inaccessible [7,16]. These challenges drove preference for longer-duration products and for biodegradable implants, which require fewer visits.

#### Injection Fear

A common barrier to LAI acceptability was dislike or fear of needles. The current formulation of cabotegravir is injected in the buttock, but both males and females in South Africa tended to dislike this location for fear of discomfort especially while sitting [23,27]. Some young women were uncomfortable having to disrobe to receive an injection in the buttocks [27].

#### Side effects

Fear of real or perceived side effects was a salient factor influencing preference, especially for younger participants–about half of male and female youth in a study in Malawi reported being unwilling to use PrEP if they experienced side effects. Pain at injection site was the most frequently mentioned concern of LAIs, although youth in qualitative interviews stated that efficacy was a more significant factor even if the formulation was expected to be painful [7,28]. Similarly, pain upon insertion was a concern regarding implants, but participants generally felt that the benefits of an effective and long-lasting PrEP product outweighed the potential for pain. Qualitative interviews indicated a preference for a flexible versus a stiff implant for increased comfort.

Pain or discomfort also influenced acceptability of the DVR, especially discomfort during intercourse. Among pregnant and postpartum women in South Africa and Kenya, most of the participants who preferred oral PrEP over a vaginal ring did so on account of concern for physical discomfort [12]. In another study conducted among FSWs in Tanzania, participants expressed concerns about infertility associated with the DVR [26]. Conversely, in two studies conducted in Kenya and in South Africa, Uganda, and Zimbabwe, participants appreciated the reversibility of the DVR and implants, which could be removed if side effects arose [11,24].

#### Oral on-demand

Three studies conducted in South Africa, Uganda, and Zimbabwe assessed preference for oral on-demand PrEP compared to daily use, and one study in Eswatini, Kenya, and South Africa compared on-demand PrEP to a long-acting product. Overall, 60% of male youth [32] and 65% of youth MSM [5] preferred on-demand PrEP over daily oral tablets, with older youth tending to have greater preference for on-demand dosing. Having more frequent sexual intercourse was associated with a lower preference for on-demand PrEP and greater preference for daily oral PrEP. Participants who preferred on-demand PrEP cited not liking daily dosing, not considering oneself at frequent enough risk to warrant taking a daily pill, stigma, and pill burden.

Conversely, those who preferred daily use cited desire for continuous and/or improved protection and comparative ease of use by dosing every day instead of having to remember to take it before sex. One study examined the effect of post-traumatic stress disorder (PTSD) on preference in SSA youth and found no significant association between PTSD symptoms and a preference for on-demand versus daily PrEP, with a 61% vs 51% preference for on-demand dosing in those with and without PTSD symptoms, respectively [33]. South African men found oral on-demand, LAIs, and implants similarly acceptable, but if only one choice was available, they preferred a once-monthly pill (32%), six-month LAIs (28%) or implants (20%) over on-demand (2%) or a two-month injectable (5%) [34].

## Discussion

This scoping review evaluated preferences and acceptability for various PrEP products across populations in SSA. Overall, we found high acceptability of LA PrEP across participant demographics and geographic region, suggesting that LA modalities can expand PrEP coverage among persons at HIV risk. The primary factors driving participant preferences for LA PrEP were efficacy, duration and discretion, which most participants felt were superior in LA products compared to oral PrEP. Overall, long-acting injectables were most preferred over the other LA products evaluated (implants and the DVR). However, we identified heterogeneity in preferences among subgroups which suggests a variety of choice will likely be needed to optimize coverage and impact of HIV prevention. This can be paralleled with the observed increase in contraceptive use with the availability of more contraceptive methods in a health system [35].

A previously published review assessed values and preferences for long-acting injectable PrEP; however, most studies included were published on or before October 2021, before regulatory approval of the first long-acting ARV for use as PrEP, CAB-LA [36]. Authors found broad interest in and overall preference for LAIs and highlighted the perceived benefits of discretion and less frequent dosing. Our review adds to the literature by evaluating other long-acting modalities in addition to LAIs and including additional publications from two years after the introduction of injectable PrEP. Further, we focus on Sub-Saharan Africa, as the region with the largest HIV burden globally as well as the most widespread rollout of PrEP worldwide. In regions of SSA with high HIV prevalence, the majority of transmission occurs through heterosexual mixing. Additionally, higher rates of oral PrEP use can influence attitudes among the general population regarding HIV prevention. One example is parental attitudes toward provisioning PrEP for their children. An overwhelming majority of caregivers of adolescents in Malawi (87%) and in South Africa (85%) expressed desire for their children to take PrEP [37,38]. This can be contrasted with a study in the American south in which parents of LGBTQ adolescents, though generally positive about PrEP, expressed relatively low intention for their children to take it [39].

Interestingly, the dapivirine ring, which was designed for use in low-resource health systems as in SSA, was a less popular choice than other long-acting options even among women with experience using DVR [7]; this was largely due to perceived side effects, especially pain during intercourse, and concern about indiscretion or an impact on the male partner’s pleasure during intercourse. One study within a DVR clinical trial found overwhelming acceptability of the ring after 28 weeks of follow-up, which suggested that the DVR could be more acceptable after experience with use [11]. However, the preferences of women choosing to participate in a DVR clinical trial may not be representative of the general population. That said, Mataboge et. al. [16] highlight focus group discussions among women [40] and their male partners [41] that suggest that while a considerable proportion of partners notice the ring during sex, the impact on sexual pleasure for both partners is minimal and in some cases positive.

Across studies, the most commonly reported concern about long-acting PrEP was potential side effects. However, empiric data shows that actual side effects were generally less frequent or severe than participants anticipated. Side effects of antivirals for PrEP are typically mild and of short-term duration, yet about half of youth participants in a study in Malawi stated they would not consider using PrEP if there were side effects [38]. Similarly, some women in interviews who had experienced side effects from injectable hormonal contraception worried about similar effects from injectable PrEP [16], although these side effects have not been observed [42]. Health communication that assuages these fears may influence the choices of users who are deterred by side effects from their otherwise preferred formulation.

Pregnant and breastfeeding women were especially concerned about side effects that could harm the fetus or infant, with some expressing fear of miscarriage or birth defects due to PrEP use [25]. Though oral PrEP is widely understood to be safe during pregnancy [43,44], other long-acting formulations have been slow to establish similar safety profiles [45]. As these data emerge, it will be imperative to educate pregnant and breastfeeding populations on the safety of these formulations to enable them to make informed decisions regarding PrEP.

Only one study compared oral on-demand PrEP with a long-acting formulation, which found similar acceptability between on-demand and long-acting products but a distinct preference for long-acting products over on-demand dosing [34]. Since participants frequently preferred on-demand over daily dosing, and because the schedule may have similar patient advantages to long-acting formulations (comparable efficacy, longer dosing frequency, increased discretion, etc.), more research is warranted in order to understand alternative product preference.

One study assessed religious background and found preference differences, namely a much greater preference for implants among Muslims compared to Christians, adherents of African indigenous religions, and non-religious participants [17]. Factors driving these preferences were not examined. Similarly, ethnicity was generally captured only on the national level in the studies included. For a region as ethnically and religiously diverse as Sub-Saharan Africa, these and other markers of cultural identity may be associated with different values surrounding sexuality and HIV prevention, and could aid local health authorities to provide the most culturally appropriate products for their communities.

Our review highlighted several gaps in existing studies. Studies on preferences of transgender women in Sub-Saharan Africa were scarce despite their having up to a 13 times higher HIV risk than that of the general population globally [46]. We identified only one study evaluating preferences in this priority population [17]. Transgender women are often grouped with MSM in preference studies, yet their preferences were distinct in the one study identified, in which transgender women tended preferred implants while MSM preferred LAIs. This highlights the importance of assessing heterogeneity in preferences across demographics as preferences of cisgender women or from other sexual minority individuals assigned male at birth do not necessarily align with those of transgender women.

We also raised a concern around generalizability for PrEP-experienced participants and especially individuals participating in clinical trials of PrEP formulations. Overall, eight studies were conducted among clinical trial participants; several preference studies were conducted during trial follow-up visits, therefore only participants who continued PrEP were included and preferences of those lost to follow up were not assessed. Findings from these studies may not be generalizable to those who are not currently taking PrEP or have significant difficulty with adherence. Individuals who may experience challenges using oral PrEP due to stigma, discretion, difficulty attending frequent refill visits, or pill burden are poorly represented in PrEP clinical trials, yet they are likely the primary target population for uptake of new PrEP modalities, as current oral PrEP coverage is low. Further, individuals participating in PrEP studies may differ from the general population in that they may be more interested in the PrEP modality evaluated in the study in which they are participating. For example, in a study that assessed preferences among participants in the DVR efficacy trial (MTN-020/ASPIRE), 94% of participants selected the vaginal ring as their most preferred LA PrEP product [11]. However, in the TRIO study, in which women were assigned to use all three of injectable, oral, and ring formulations, only 12% of individuals most preferred the vaginal ring [47]. Future studies should consider investigating PrEP preferences among oral PrEP-naïve individuals not participating in PrEP studies.

Finally, the results of this review are limited by the hypothetical nature of many of the studies. Cabotegravir is the only LAI currently available, and 11 of 25 studies assessing LAIs were published before cabotegravir was approved for use as PrEP. Similarly, implantable PrEP is still in development. In both cases experience with injectable or implantable contraceptives can be a useful proxy for experience with that modality as a PrEP product [14]. However, participants analogizing injectables and implants with hormonal contraceptives often feared side effects similar to such contraceptives, even though the side effect profile for ARVs is milder than that for contraceptive hormones. Further research will be needed to understand in-practice preference for long-acting modalities outside of clinical trials as LA PrEP becomes widely available.

### Conclusion

Long-acting PrEP formulations are highly acceptable across demographics in SSA and can increase PrEP coverage to meet global targets for HIV prevention. Overall, injectable PrEP was most preferred followed by biodegradable implants, with product duration playing the most salient role in preferences. The intravaginal ring was the least preferred LA product but still more preferred than daily oral PrEP. There was significant interest in on-demand oral dosing among lower-risk participants with less frequent exposures. Further research will be needed to understand enacted preference as these and other long-acting modalities become available to patients.

## Data Availability

All data produced in the present work are contained in the manuscript.

## Authors’ contributions

All authors contributed to developing the analysis plan, interpreting results, and commenting on manuscript drafts. MS conceived of the analysis. BP, AS, and SC conducted the literature review and synthesis. BP wrote the first draft of the manuscript. All authors had full access to the data in the study and had final responsibility for the decision to submit for publication. All authors read and approved the final manuscript.

## Acknowledgements

We thank Nicole Young, Shirley Chen, and Geoff Garnett at the Bill and Melinda Gates Foundation for their support and funding of this analysis.

## Supplement

Table S1. Search strings

Table S2. Quality assessment checklist

