## Supplemental Material for "User Preferences on Long-Acting Pre-Exposure Prophylaxis for HIV Prevention in Sub-Saharan Africa: A Scoping Review"

Table S1. Search strings.

| **Search String** | **# Total results** | **# Relevant results** |
| --- | --- | --- |
| (PrEP) AND (Long-acting) AND (Discrete Choice) | 12 | 4 |
| (PrEP) AND (Long-acting) AND (preference) | 92 | 21 |
| (PrEP) AND (preference) AND (Africa) | 123 | 28 |
| (PrEP) AND (on demand) AND (preference) | 59 | 7 |

Table S2. Quality assessment checklist.

| **Author et al. (Year)** | **Generalizability to target population** | **Participant acceptance rate** | **PrEP-experienced sample?** |
| --- | --- | --- | --- |
| Mack et al. (2014) | No major concerns | Not reported | No |
| Luecke et al. (2016) | Participants recruited from earlier VOICE clinical trial | Not reported | Yes; clinical trial |
| Van der Straten et al. (2017) | Participants from clinical trial | Not reported | Yes; clinical trial |
| Krogstad et al. (2018) | Some participants were former iPrevent or TRIO study participants | Not reported | 24 of 105 participants were contraceptive implant-experienced; 13 were PrEP LAI-experienced |
| Siedner et al. (2018) | No major concerns | Not reported | No |
| Van der Straten et al. (2018) | Participants from clinical trial | 277/449 (61.7%) | Yes; clinical trial |
| Cheng et al. (2019) | No major concerns | Not reported | Not reported |
| Harling et al. (2019) | Demographic limited to female bar workers | 66/66 (100.0%) | No; few end-users were aware of PrEP |
| Krogstad et al. (2019) | No major concerns, however, end-users were not sampled | Not reported | NA; study did not examine end-users |
| Montgomery et al. (2019) | Recruited from past clinical trial participants | Not reported | Yes. Purposively sampled experienced users |
| Tolley et al. (2019) | Participants from clinical trial | Not reported | Yes; clinical trial |
| Kidman et al. (2020) | No major concerns; nested within the Malawi Longitudinal Study of Families and Health | 2089/3317 (63.0%) | No |
| Laher et al. (2020) | Participants recruited from the HVTN 702 vaccine trial | 38/71 (53.5%) | Some, proportion not reported |
| Minnis et al. (2020) | No major concerns | Not reported | No, though most girls had used contraceptive methods |
| Van der Straten et al. (2020) | No major concerns | Not reported | 5 of 128 participants had used PrEP |
| Montgomery et al. (2021) | No major concerns; purposively sampled clinical reserach-naive participants | Not reported | Some, proportion not reported |
| Ngure et al. (2021) | Follow-up from MPYA study | 642/934 (68.7%) | Yes; clinical trial |
| Beckham et al. (2022) | No major concerns; participants were recruited from Project Shikamana trial, which evaluated a community HIV response intervention, not a PrEP product | Not reported | No; few end-users were aware of PrEP |
| Brown et al. (2022) | No major concerns | Not reported | No; few end-users were aware of PrEP |
| Dietrich et al. (2022) | No major concerns | Not reported | No |
| Little et al. (2022) | No major concerns; sampled through respondent-driven sampling | Not reported | 20% of participants had used PrEP, and 33% had heard of it |
| Mayanja et al. (2022) | Participants from cohort study | 285/532 (53.6%) | No |
| Ogunbajo et al. (2022) | No major concerns | Not reported | 15% were PrEP-experienced |
| Webb et al. (2022) | No major concerns | Not reported | Not reported |
| Bailey et al. (2023) | No major concerns | Not reported | 65% had ever taken PrEP; 31% were taking PrEP at the time of the study |
| Jansen van Vuuren et al. (2023) | Participants from cohort study | 425/635 (66.9%) | No |
| Kakande et al. (2023) | Participants from clinical trial | 33/37 (89.2%) | Yes; clinical trial |
| Little et al. (2023) | Participants were recruited through market research databases and averaged higher in education and wealth status than the general population | Not reported | 16% had used PrEP |
| Mataboge et al. (2022) | Female participants were recruited from Project PrEP and had consented to participate in research | Not reported | 34% had ever used PrEP. Purposively sampled users and non-users |
| Ngure et al. (2023) | Participants from clinical trial | Not reported | Yes; clinical trial |
| Tran et al. (2023) | Product preferences for ART assessed among HIV+ participants | Not reported | HIV+ participants on ART |
| Wara et al. (2023) | Participants from cohort study and clinical trial | 394/424 (92.9%) | 75% had used PrEP in last 30 days. |
| Mthimkhulu et al. (2024) | No major concerns | Not reported | 24% had used PrEP |
| Gates Foundation [not published] | Sampling method not described | Not reported | Not reported |
| Were [not yet published] | No major concerns | Not reported | Most had used PrEP; 29% declined to use |
